# The straight and narrow: a game theory model of broad- and narrow-spectrum empiric antibiotic therapy

**DOI:** 10.1101/2023.02.15.23285947

**Authors:** Maya Diamant, Uri Obolski

## Abstract

Physicians prescribe empiric antibiotic treatment when definitive knowledge of the pathogen causing an infection is lacking. The options of empiric treatment can be largely divided into broad- and narrow-spectrum antibiotics. Prescribing a broad-spectrum antibiotic increases the chances of covering the causative pathogen, and hence benefits the current patient’s recovery. However, prescription of broad-spectrum antibiotics also accelerates the expansion of antibiotic resistance, potentially harming future patients. We analyze the social dilemma using game theory. In our game model, physicians choose between prescribing broad and narrow-spectrum antibiotics to their patients. Their decisions rely on the probability of an infection by a resistant pathogen before definitive laboratory results are available. We prove that whenever the equilibrium strategies differ from the socially optimal policy, the deviation is always towards a more excessive use of the broad-spectrum antibiotic. We further show that if prescribing broad-spectrum antibiotics only to patients with a high probability of resistant infection is the socially optimal policy, then decentralization of the decision making may make this policy individually irrational, and thus sabotage its implementation. We discuss the importance of improving the probabilistic information available to the physician and promoting centralized decision making.

## 1 Introduction

The emergence and spread of antibiotic-resistant bacteria is a significant public health problem world-wide [1], as antibiotic resistant infections increase mortality rates and treatment costs [2]. A proposed possibility to alleviate this problem is to develop novel, broad-spectrum antibiotic drugs. However, developing and producing new antibiotic drugs is a prolonged and costly process, and the production rate of new antibiotics has substantially decreased over the years [3, 4]. Furthermore, high consumption of antibiotics accelerates the development of resistance, quickly rendering new drugs less effective [5, 6]. Therefore, new-generation broad-spectrum antibiotic drugs are a scarce resource [7], and should be administered with utmost caution.

This attitude is at odds with empiric antibiotic therapy - the commencement of treatment without definitive diagnosis of the causative pathogen. Physicians prescribing empiric therapy thus face a dilemma between administering the patients efficient initial empirical antibiotic treatment and the emerging need in reducing the use of broad-spectrum drugs. Empirical treatment that ex-post matches the causative pathogen, is highly important, as it reduces the mortality rate due to bacterial infections [8, 9]. Naturally, a broad-spectrum drug has much higher probability for such a match, compared to a narrow-spectrum drug. Thus, on one hand, the best interest of the individual patient may very well be receiving broad-spectrum antibiotics. On the other hand, as mentioned above, there is a social and ecological utility in minimizing the use of broad-spectrum antibiotics. Hence, the best interest of the entire (potential) patients’ population might be referring the broad-spectrum drug as a “weapon of last resort”, administering it only to those patients who most likely need it.

This problem is considered a social dilemma, whose mitigation deserves awareness and collective action [10, 11]. A few studies have addressed this problem using economic models, aiming to incentivize investments in hospital infection control and development of new antibiotics [12, 13]. Other studies have analyzed the clinical aspects of this decision making problem. For example, Parra-Rodriguez et al. [14] elaborated on the information available to the physician throughout the decision process, and the possible role of decision support systems. Barash et al. [15] emphasized the importance of performing proper and non-biased analytical reasoning by the physician.

This conflict of interests is especially amenable for a game-theoretic analysis. As such, previous game-theoretic studies have focused on related problems in antibiotic prescription, mostly pertaining to empiric therapy of viral vs bacterial infections. For example, Evolutionary game theory was used to model definitive antibiotic treatment [16], demonstrating that fast social learning may help curb the overuse of antibiotics. In addition, two behavioural game theory studies showed that providing patients with social information about other people’s antibiotic intake[17] and eliciting empathy toward future patients[18] may help to reduce antibiotic overuse in mild diseases. However, these studies did not consider uncertainty regarding patient diagnoses, which is a key component in empiric therapy. On the other hand, when analyzing the problem of empiric antibiotic treatment using game theory, Colman et al. [19] showed that if physicians ignore long-term implications of antibiotic usage and base their clinical decision only on the prior probability of bacterial infection, then administering an empiric antibiotic treatment is a dominant strategy. In our previous study [20], each physician did have long-term consideration regarding her own future patients, and the clinical decision was based on the differential information regarding the posterior probability of bacterial infection in each patient. We showed that in equilibrium, the threshold probability of antibiotic treatment is lower than the socially optimal one. Nonetheless, game theory has not yet been used to address the problem of choosing between broad and narrow-spectrum antibiotics.

In the settings of this study, each patient is infected by a resistant (*R*) or a susceptible 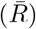 bacterial strain. Two types of antibiotic treatments are available: narrow- (*N*), and broad-spectrum (*B*). A susceptible infection is successfully cleared by both types of antibiotics, whereas a resistant infection has a substantially higher probability of clearance under treatment with the broad-spectrum antibiotic. The physician, an agent acting on behalf of the patient, must decide which empiric treatment should be administered. Her decision relies on partial information about the patient: the likelihood of a resistant bacterial infection in a given patient before culture results are available (derived from prior knowledge or clinical manifestations).

We assume that each physician is totally and equally committed to each of her patients’ health. Therefore the monetary cost of treatment is not a factor in her decision, and she aims to maximize the cumulative utility of all of her own patients over time, given the behaviour of the other physicians. By contrast, a social planner aims to maximize the cumulative utility of all the patients (regardless of their physician) over time.

The success of the treatment depends on the drugs’ effectiveness in clearing the infection – the probability that the infecting pathogen is susceptible to the drug. The broad-spectrum antibiotic is defined as having a higher initial effectiveness. However, each use of an antibiotic increases the future resistance to it, and thus decreases its effectiveness [21, 22, 23]. In principle, each antibiotic treatment also exposes the patient to risks such as treatment side effects or increased likelihood of developing other, antibiotic resistant bacterial infections [24, 25, 26, 27]. Nonetheless, for simplicity, and since these risks are low relative to the immediate risk of not clearing the infection, we neglect these them. We start by specifying the details of the game model. We then consider the use of broad-spectrum antibiotics under a socially optimal policy, which takes into consideration the entire patient population. This policy is compared to the level of broad-spectrum antibiotic use in the subgame-perfect equilibrium, where each physician considers only the utility of her own patients. We end by discussing the meaning of these results, emphasizing some practical aspects.

## 2 Model

We consider a game of *n* players (physicians) denoted *i ∈*{1, 2, …, *n*}. The game lasts *T* + 1 periods, *t* = 0, 1, …, *T*, in which each of the players treats a single patient. The socially optimal policy will be derived by assuming that one physician treats all the patients.

As mentioned, each patient is either infected by a resistant (*R*) or susceptible 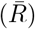 bacterial strain. The true diagnosis of resistant/susceptible infection of the patient (“H-state”) is unknown to the players, who only have a probabilistic information based on symptoms and immediate diagnostic tests. For simplicity, a patient can either have a specific set of characteristics or not have it. For example, these characteristics might be previous use of antibiotics, indication from rapid diagnostic tools and so forth [23, 27, 28]. We will refer to these as a patient’s “signal”, which is either “high” (*H* with probability *q*_*H*_) or “low” (*L* with probability *q*_*L*_ = (1 − *q*_*H*_)). The likelihood of resistant infection given the signal *H* is *p*_*H*_, and the likelihood given *L* is *p*_*L*_, with *p*_*H*_ > *p*_*L*_. Since the current model deals with a limited horizon (the number of periods is relatively small), we assume that *q*_*H*_ is fixed.

Each player knows only the signal of her own patient. Based on this information, each player may choose one of two possible actions: to either administer broad-spectrum (*B*) or narrow-spectrum (*N*) antibiotic treatment.

We further assume that (a) due to the risk of a resistant bacterial infection, treating it is always preferred to not treating it; and (b) having a susceptible bacterial infection is preferred to having a resistant one, regardless of whether the treatment is with a broad or narrow drug. Thus, we can define three levels of utility: *r*_1_, the utility (or survival rate) of patients with a resistant infection given inappropriate empirical treatment; *r*_2_, the utility of patients with a resistant infection given appropriate empirical treatment; and *r*_3_, the utility patients with a susceptible infection given appropriate empirical treatment, where *r*_3_ > *r*_2_ > *r*_1_ (note that the assumption regarding *r*_3_ will not be needed later on in our complete model, as explained in section 2.3).

This decision problem is illustrated for a single patient in the decision tree shown in figure 1, and the definitions of all relevant variables are summarized in table 1.

**Figure 1:**
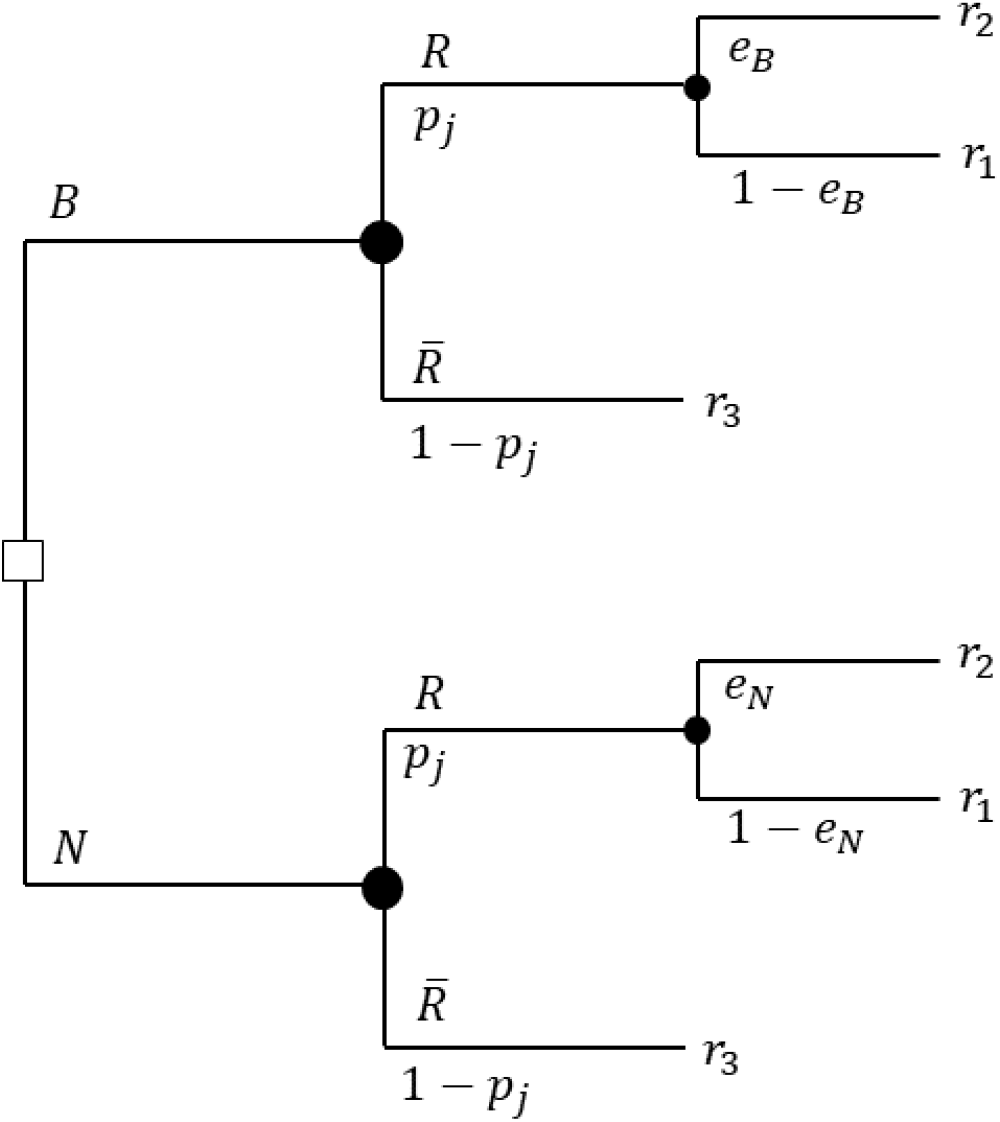
The decision tree of the single-patient problem, given signal *p*_*j*_. The tree root starts with a decision (white square) of whether to treat with a broad (*B*) or narrow (*N*) spectrum drug. Then, the probabilities of a patient having a resistant (*R*) or susceptible 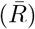 bacterial infection, and the relevant drug’s effectiveness in clearing the infection (*e*_*B*_, *e*_*N*_). The definitions of relevant variables are summarized in table 1.

**Table 1:** Definitions of the variables of the single-patient problem

|  |  |
| --- | --- |
| $i \in \{1, 2, \dots, n\}$ | Players (physicians / hospitals / countries) |
| $j \in \{H, L\}$ | Patient signal (set of symptoms, high/low probability of infection by a resistant bacteria) |
| $p_j$ | Probability of resistant bacteria given signal $j$, $j \in \{H, L\}$ |
| $q_j$ | The probability that the patient has signal $j$, $j \in \{H, L\}$ |
| $B, N$ | Broad/narrow-spectrum antibiotic drug |
| $r_1$ | Utility (or survival rate) of resistant infection given inappropriate empirical treatment |
| $r_2$ | Utility (or survival rate) of resistant infection given appropriate empirical treatment |
| $r_3$ | Utility (or survival rate) of susceptible infection given appropriate empirical treatment |

### 2.1 Effectiveness Depletion

Each use of an antibiotic drug depletes its future effectiveness due to antibiotic resistance [21, 22, 23, 27]. We assume that each use of a drug in the current period detracts its average effectiveness in the following period by a constant depletion effect, which may vary between drugs [26]. Formally, let 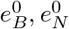 be the initial effectiveness levels of drugs *B, N* respectively, where 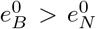 (i.e., the broad-spectrum drug is more likely to be effective). We denote *α*_*B*_ (*α*_*N*_) the marginal depletion effect of individual usage of drug *B* (*N*) on drug effectiveness.

We assume that use of *N* does not deplete the effectiveness of *B*. However, due to cross-resistance [29], the use of *B* can reduce the effectiveness of *N*. We therefore define *I ∈* [0, 1] as the effectiveness depletion independence level: (1 − *I*) is the fraction of *α*_*N*_ subtracted from 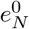 by each use of *B*. Specifically, *I* = 0 means that the broad-spectrum antibiotics “contain” the narrow ones, in the sense that each use of *B* reduces the effectiveness of both *B* and *N* (by *α*_*B*_ and *α*_*N*_ respectively); *I* = 1 means that the use of a certain drug does not induce resistance to the other drug.

The current effectiveness-state of the entire system (“E-state”) is given by the couple (*t, k*), where *t* is the current period and *k* is the number of patients who were treated with *B* in the previous periods (prior to *t*). The transition between E-states is determined by the players’ actions and the current E-state, through the effectiveness depletion dynamics. The graph in figure 2 demonstrates the possible game states and transitions for a 2-player, 3-period game.

**Figure 2:**
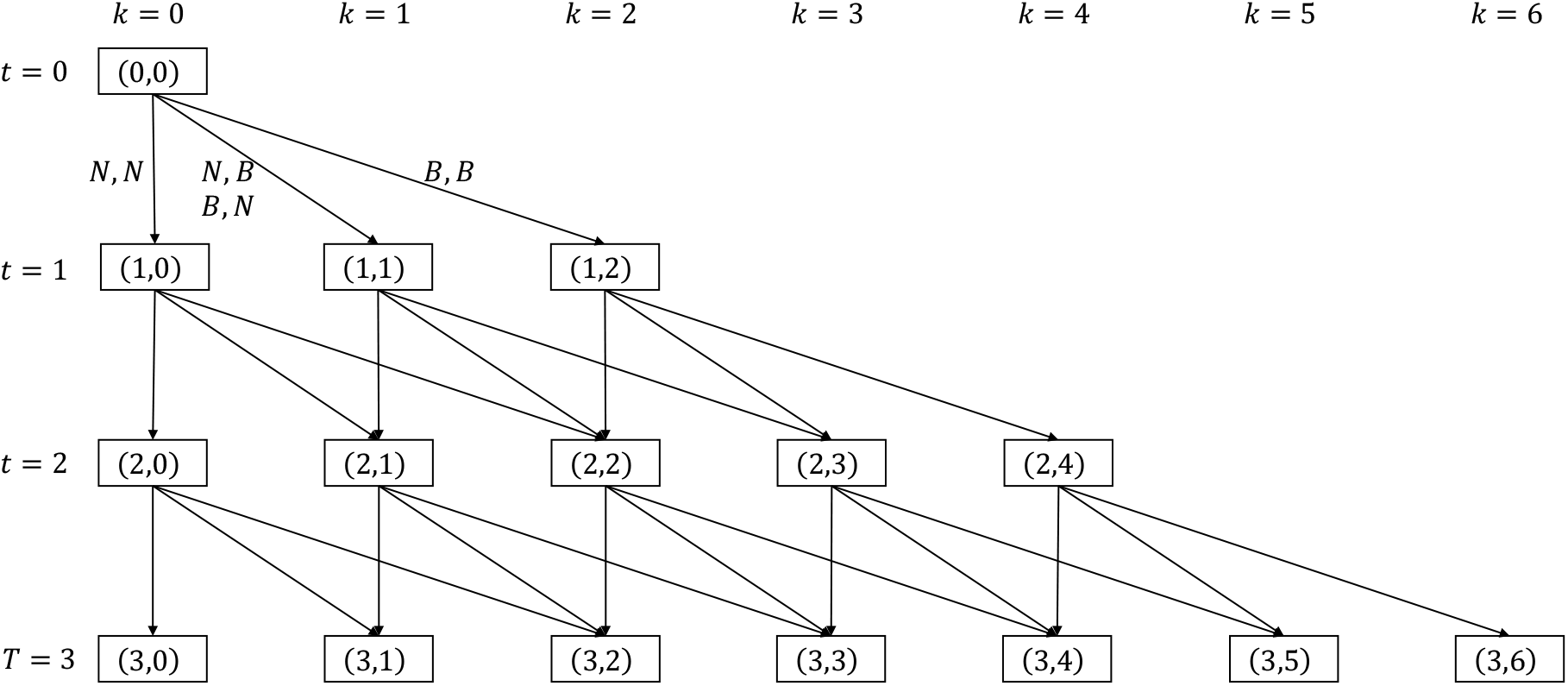
The E-states and possible transitions for a 2-player, 3-period game.

Let 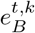 be the effectiveness of *B* at E-state (*t, k*).

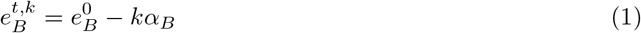

Let 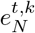 be the effectiveness of *N* at E-state (*t, k*).

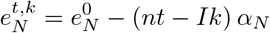

If *I* = 1 we get

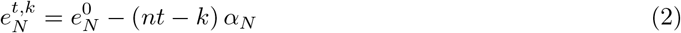

And if *I* = 0 we get

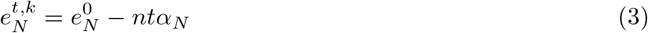

The definitions of all relevant variables are summarized in table 2.

**Table 2:** Definitions of the variables of the effectiveness depletion dynamics

|  |  |
| --- | --- |
| $e_B^0, e_N^0$ | Initial effectiveness (coverage percentage) of antibiotic drug $B/N$ ($e_B^0 > e_N^0$) |
| $\alpha_B, \alpha_N$ | The marginal effect of individual usage of drug $B/N$ on resistance development (effectiveness depletion) |
| $t = 0, 1, \dots, T$ | Time ($T + 1$ periods) |
| $k \in \{0, 1, 2, \dots, nT\}$ | Number of patients who were treated with $B$ in the previous periods. |
| $(t, k)$ | The E-state of the system |
| $I \in [0, 1]$ | Effectiveness depletion independence level. $(1 - I)$ is the fraction of $\alpha_N$ subtracted from $e_N$ by each use of $B$.<br>(Use of $N$ does not deplete $e_B$ ).<br>$I = 0 \Rightarrow$ Full inclusion<br>$I = 1 \Rightarrow$ Independence |
| $e_B^{t,k}$ | The effectiveness of $B$ at E-state $(t, k)$. $e_B^{t,k} = e_B^0 - k\alpha_B$ |
| $e_N^{t,k}$ | The effectiveness of $N$ at E-state $(t, k)$. $e_N^{t,k} = e_N^0 - (nt - Ik)\alpha_N$ |

We assume a finite game, with a horizon that is sufficiently short to maintain the assumption of constant parameters as reasonable. One implication of the short horizon is that *q*_*H*_ remains fixed for all *t*. Another main implication is that even if all patients are treated with the broad-spectrum antibiotic, its effectiveness remains higher than that of the narrow-spectrum antibiotic.

The initial conditions of the parameters are:

**Condition 1**. *Drug B remains more effective even if all patients receive it:*

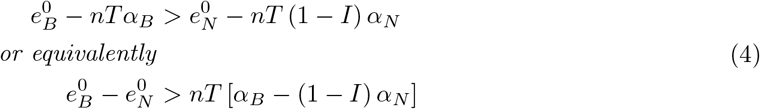

**Corollary 1**.

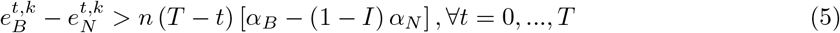

**Condition 2**. *The drug N is not completely ineffective even if all patients receive it:*

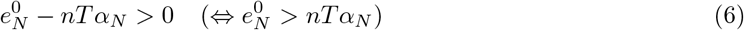

### 2.2 decision rules and strategies

A strategy of a physician is a mapping from histories (of states and actions of all the players) to actions. However, in the context of medical treatment, strategies that rely on the past behaviour of other players (both “punishments” and “rewards”) seem highly implausible. For example, it is highly unlikely that a physician will decide to prescribe broad-spectrum antibiotics to her current patient because another physician did so in the previous period. Therefore, we shall concentrate on Markovian strategies. Markovian strategies depend only on payoff-relevant variables, which in our model are the states (the E-states and the partial information about the H-states). Thus, we are interested in finding the subgame perfect equilibrium in Markovian strategies (also known as Markov perfect equilibrium -MPE).

To check whether a combination of Markovian strategies is an equilibrium, we need only to check that each player has no incentive to deviate to another Markovian strategy. This results from the fact that given any fixed stationary Markov strategy played by the other physicians, the decision problem faced by physician *i* is equivalent to a Markov decision process (MDP)[30]. Thus, a best response exists in Markov strategies, and it can be found using a maximization process of dynamic programming. Therefore, we will denote strategies as Markovian, even though we do not actually limit a physician from deviating into a non-Markov strategy.

A Markov strategy is compounded of decision rules. A decision rule of a physician determines what to do in the current E-state, given the signal that he currently observes (the physician’s information about the current H-state), and not on the history.

Let 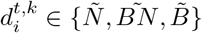 be the decision rule of player *i* in E-state (*t, k*). We will limit our discussion to the three possible “threshold decision rules”. These threshold decision rules are not only the most intuitive, it is also easy to prove that they are the most efficient.

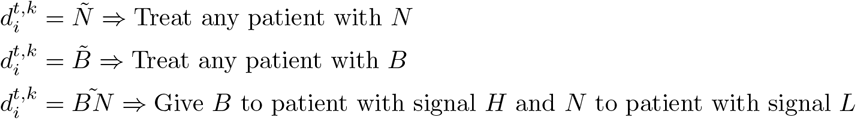

We further denote by 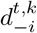 the decision rule of all the other players except for *i* in E-state (*t, k*). This paper concentrates on symmetric strategies, and thus we assume all the other player use the same decision rule at any E-state. When modeling asymmetric strategies, 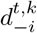 can be replaced by a (*n* − 1)- tuple: 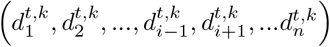. A symmetric decision rule of a social planner in E-state (*t, k*) is denoted *d*^*t,k*^.

Therefore a pure Markov strategy *s*_*i*_ of physician *i* is an array of (*T* + 1) vectors, labeled *t* = 0, 1, …, *T*. Each vector contains player *i*’s decision rules for all the possible E-states in period *t* ((*nt* + 1) decision rules):

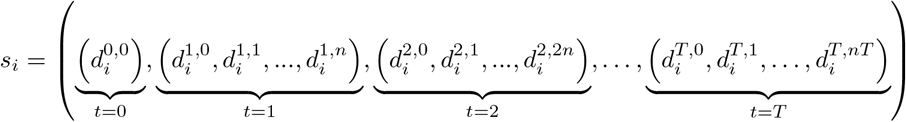

Note that if player *i* chooses strategy *s*_*i*_, not all the decision rules that it contains will necessarily be applied in the realization of the game. For example, if the current E-state is (*t, k*) and *m* > 0 physicians choose *B*, the game moves to E-state (*t* + 1, *k* + *m*), and player *i* will not implement 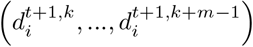. We denote by 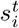 the projection of *s*_*i*_ from period *t* onward (the “tail” of the array, starting from vector *t*).

The definitions of all the variables relevant to strategy definitions and to the payoff calculations described in the following section are summarized in table 3.

**Table 3:**
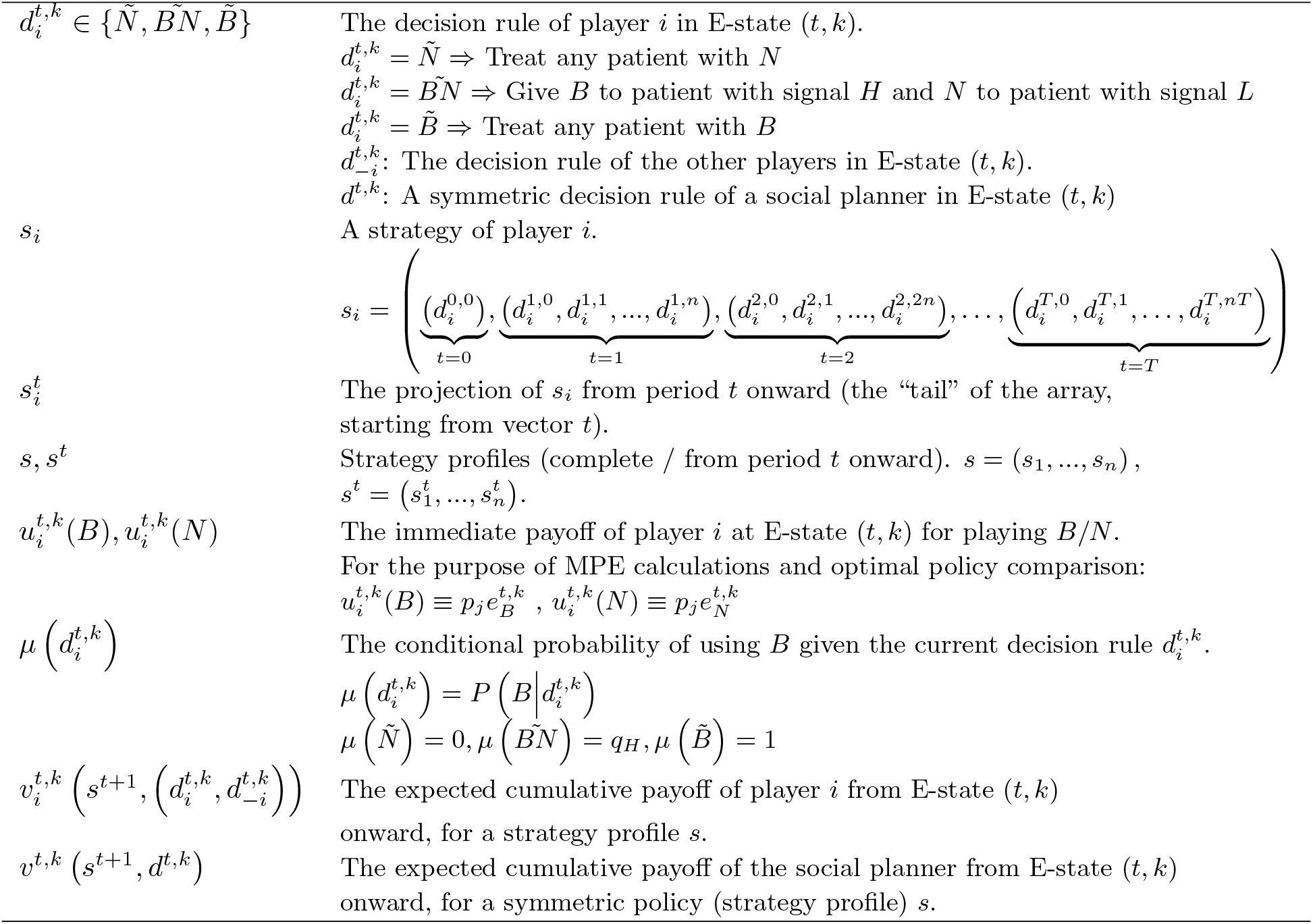
Definitions of the variables in the static decision problem

### 2.3 Payoffs Calculation

The immediate payoffs in E-state (*t, k*) are:

If player i plays *B*:

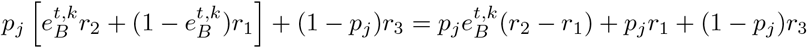

If player i plays *N* :

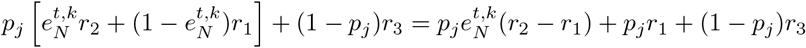

Where *j* ∈{*H, L*} is her own patient’s signal, and the other parameters are defined in table 1.

Since we are interested in comparing the the expected payoffs of different actions, we can subtract identical components from all payoffs and multiply all payoffs by a positive constant, without affecting the strategic analysis. Thus, we define the immediate payoffs in period *t* as follows:

If player i plays *B*:

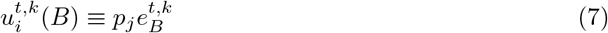

If player i plays *N* :

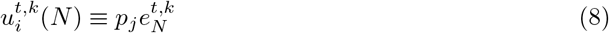

Notice that the immediate payoff of each player depends only on his own action in the current period. The mutual influence between the players is indirect and delayed, through the depletion of *e*.

Let 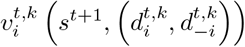 be the expected cumulative payoff of player *i* from E-state (*t, k*) onward, for a strategy profile *s*. Note that the payoff depends on the strategies of the players only from this point onward (i.e. their decisions for E-state *t, k* and for the E-states in periods *t* + 1, …, *T*), and on the distribution of the future patients’ signals.

This expected cumulative payoff 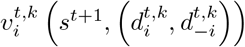 can be calculated recursively, starting from the E-states of period *T* and moving backward. Generally speaking, it equals the expected utility in the current E-state (with respect to the probability distribution of the patients’ signals) and a probabilistic transition function to E-states in the next period:

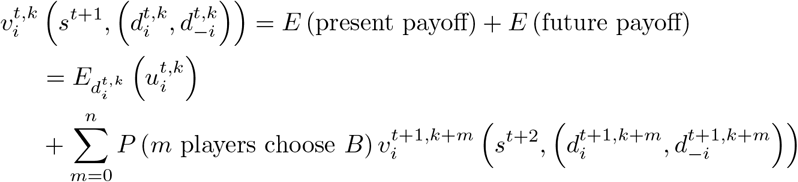

The expected present payoff depends on the current E-state, the player’s own decision rule in the current E-state and the distribution of patient signals:

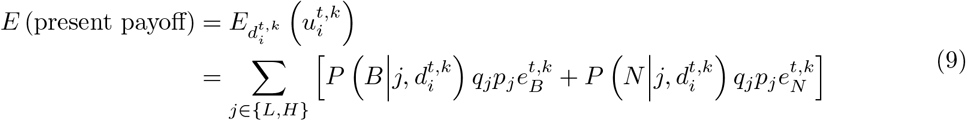

If 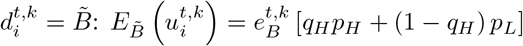

If 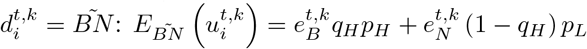

If 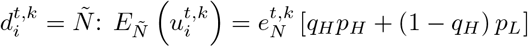

Due to (**??**), the immediate payoff from using *B* is higher than *N*, and therefore

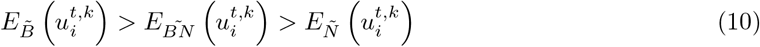

For the purpose of calculating the expected future payoff, let 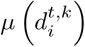 be the conditional probability of using *B* given the current decision rule 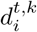.

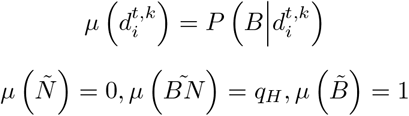

When all the players use the same decision rule in the current E-state, 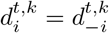, the total expected payoffs can be calculated using the following recursive equation:

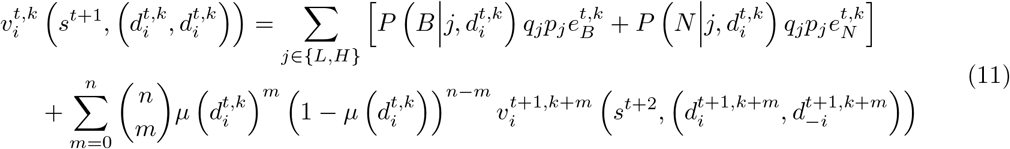

And when player *i* deviates to a different decision rule, 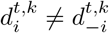, we get:

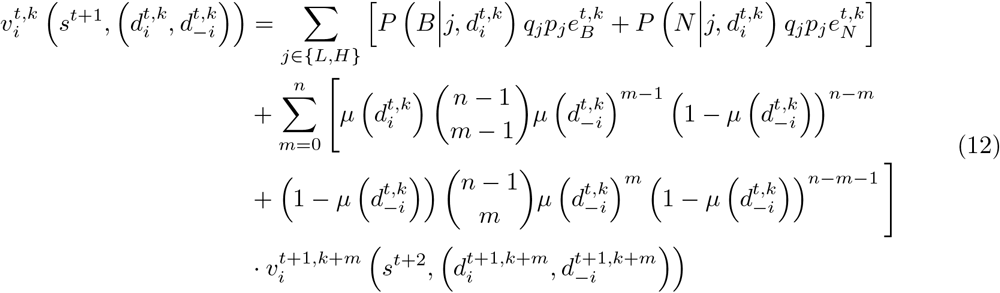

We assume the social planner uses symmetric policies. That is, it employs an identical decision rule *d*^*t,k*^ on all the current patients in E-state *t, k*. Therefor, the expected cumulative payoff of the social planner from E-state (*t, k*) onward, for a symmetric policy (strategy profile) *s* equals

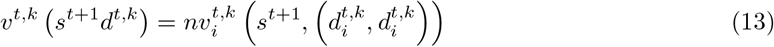

## 3 Results

The socially optimal policy and the players’ strategies in Subgame-Perfect Nash-Equilibrium in Markovian strategies (MPE) can be found through backward induction.

In the last period of the game, prescribing the more effective drug regardless of the patient’s signal is a dominant action - both for the social planner and for any rational individual player. Under condition 1, this dominant action is *B*. This claim is formalized and proven in the supplementary material (SM.4).

When continuing the backward induction process to preceding periods, finding either the MPE strategies or the socially optimal policy relies on the differences between the expected payoffs from alternative E-states onward. We focus on cases in which the medical parameters of the problem and the strategy profile of the players (or the policy of the social planner) induce the following condition:

**Condition 3** (Normal-differences condition).

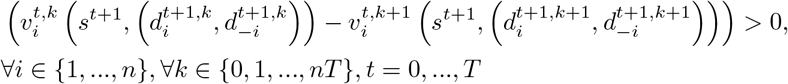

This condition implies that at any given period *t*, player *i* may gain a higher expected payoff from this point onward when having a higher effectiveness of the broad-spectrum drug.

Note that this condition is not trivial and might not apply under certain combinations of medical parameters. For instance, if *α*_*B*_ *≪α*_*N*_ then a physician may gain a higher utility by having a slightly lower effectiveness of *B* combined with much higher effectiveness of *N* (drug *N* becomes the scarce resource). However, these circumstances are extreme and thus are of less interest.

In addition, condition 3 may not apply due to an arbitrary or “weird” behaviour of one or more players in future periods (or a sub-optimal policy of a social planner). However, we assume all players are rational and thus utility maximizers. It can be easily shown, for example, that if *I* = 0 the condition applies under socially optimal policy for any medical parameters.

Therefore, we refer to this condition as “normal”, and our analyses and discussion concentrates on scenarios where it holds.

Our first theorem states a general major result:

Under the Normal-differences condition, *if the decision rule* 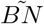 *at any specific game-tree node (E-state) is part of an MPE, then it is socially optimal at this E-state as well; however, if it is socially optimal at a certain E-state, an individual player may have an incentive to deviate towards an excessive use of B*.

For formulation and proof of Theorem 1 see the supplementary material (SM.3).

This theorem entails that whenever the MPE strategies differ from the socially optimal policy - the deviation is always towards a more excessive use (i.e. a faster depletion) of the broad-spectrum drug.

**Corollary 2**. *The expected administration of the broad-spectrum drug, at each game-tree node (E-state), and thus in the entire game, is at least as high under the MPE as under the socially optimal policy*.

An interesting special case is the fixed symmetric policy (strategy profile) of “treating a high-signal patient with *B* and treating a low-signal patient with *N* “ (except for the last period, where any patient is treated with *B*). This policy reflects the perception that *B* is a “weapon of last resort”. We will refer to it as the “signal-based” policy (strategy profile).

Under the signal-based policy, the difference between the expected payoffs from two adjacent E-states onwards can be presented explicitly:

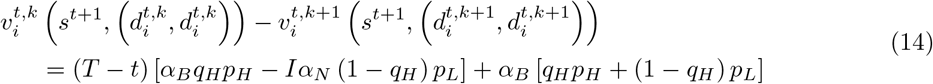

For formulation and proof see claim 2 on the supplementary material (SM.5).

Thus, if we ignore the effect of the last period, we get that the Normal-differences condition will apply only if

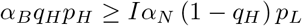

or equivalently

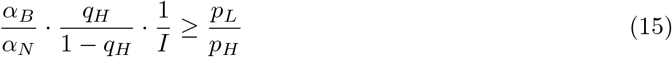

Since 0 < *α*_*B*_, *α*_*N*_, *q*_*H*_, *p*_*H*_, *p*_*L*_ < 1, equation (15) is always true for *I* = 0 (full inclusion, i.e. any use of *B* depletes *N* by *α*_*N*_), and for a wide range of other parameters. The most limited case is when *I* = 1 (full independence, the use of *B* does not deplete *N*). Figure 3 illustrates the possible combinations of parameter ratios for the cases of *I* = 1 and *I* = 0.1. For a given 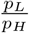 ratio, the Normal-differences condition applies for parameter combinations (of 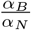 and *q*_*H*_) that are above the relevant 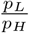 curve.

**Figure 3:**
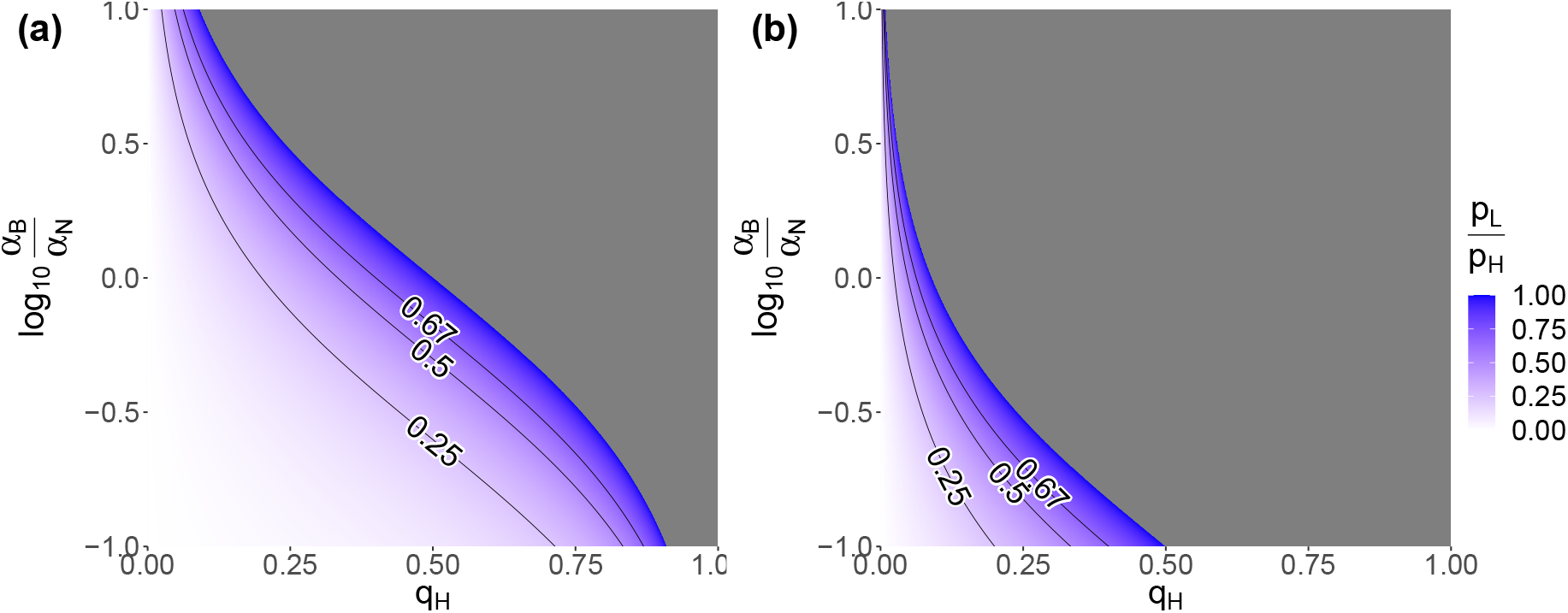
The normal-differences condition for the policy of treating a high-signal patient with *B* and treating a low-signal patient with *N*. The horizontal axis presents the possible values of the high-signal frequency *q*_*H*_ and the vertical axis presents the depletion effects ratio in log-scale, log_10_ 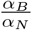. Colours represent the the values of 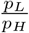 needed to satisfy the normal condition: 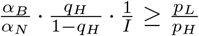. In the grey area, the left-hand side of the above expression is greater than 1 and thus the condition necessarily applies (since 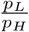< 1 by definition). In (a) *I* = 1, i.e. each use of *B* only reduces the effectiveness of *B* (by *α*_*B*_); in (b) *I* = 0.1, i.e. each use of *B* reduces the effectiveness of *B* by *α*_*B*_ and of *N* by 0.9*α*_*N*_.

It demonstrates that the space of possible parameter combinations for which condition 3 holds under the “treating a high-signal patient with *B* and treating a low-signal patient with *N* “ policy grows as *I* decreases.

Equation (14) enables us to simplify the conditions for MPE and socially optimal policy underlying Theorem 1, and introduce Theorem 2:

> *If condition 3 holds, then if the “signal-based policy” is socially optimal, individual players may have an incentive to deviate towards an excessive use of B. Furthermore, if this policy is not socially optimal for a given combination of medical parameters (e*_*B*_, *e*_*N*_, *α*_*B*_, *α*_*N*_, *q*_*H*_, *p*_*H*_, *p*_*L*_, *I) when n* = 1, *it may become socially optimal as the number of patients grows, but this strategy-profile will never be an MPE*.

For a formal formulation and proof of Theorem 2 see the supplementary material (SM.8).

This theorem has a significant practical meaning: the more decentralized the decision making is, the more we can expect excessive administration of the broad-spectrum drug. The game-theoretic intuition is that when a single player unilaterally deviates towards overusing the broad-spectrum drug she takes the strategies of the other players as given and only considers the future consequences for her own patients, while a centralized decision maker (social planner) considers the overall implications.

## 4 Discussion

We have shown that rational considerations may lead individual physicians to deviate from the socially optimal policy towards over-prescription of broad-spectrum antibiotics. In real-life settings, the socially optimal policy, which takes into consideration the impact of antibiotic treatment on future patients, is reflected in medical guidelines for empiric antibiotic therapy (see for example [31, 32, 33]). Previous works are in agreement that physicians often fail to adhere to such guidelines, resulting a much higher prescription rate of antimicrobial therapy generally, and particularly of broad-spectrum antibiotics[34, 35]. Our analysis suggests that this behaviour should not be dismissed as a lack of knowledge or an error in judgement, but may alternatively be interpreted as a deliberate, rational strategic behaviour. This interpretation is consistent with previous works’ findings that deviation from the guidelines towards over-prescription of antibiotics in general, and specifically for broad-spectrum antibiotics, is more prevalent among experienced physicians and experts[35, 36].

The main focus of this work is the strategic decision making of physicians, when facing multi-dimensional uncertainty: uncertainty regarding the diagnosis (type of causative pathogen) of their present patient, the diagnoses of other physicians’ present patients, the diagnoses of future patients, and uncertainty regarding the present and future decisions of the other physicians. To handle the complexity of this multi-player incomplete-information stochastic game analytically, we relied on a few simplifying assumptions. We assumed that the information signal presented by patients is dichotomous, and that each physician treats exactly one patient per period. Although future work may relax these assumptions into more generalized settings, we believe that these will not affect the nature of our conclusions, because the essence of the problem will not vanish: As long as each physician considers the future consequences of antibiotic prescription only for her own patients, and not the overall implications, she may have an incentive to deviate from the socially optimal policy. In addition, we assumed that the frequency of the high-signal symptom, *q*_*H*_ is fixed throughout the game. In particular, a fixed *q*_*H*_ is independent of the current level of effectiveness. Future research may consider a long-term dependence between these variables (i.e. the probability of being infected by a resistant bacteria increases as drug effectiveness declines) and thus be relevant to modelling longer game horizons.

Obviously, the information available to physicians and their ability to make probabilistic reasoning play a major role in their decisions [15]. Physicians’ decisions rely on the probability of a resistant bacterial infection before definitive laboratory results are available. In our setting, this probability is either relatively high, *p*_*H*_, or low, *p*_*L*_, depending on an information signal, which is based on patient’s medical symptoms, clinical settings, personal data, or the results of immediate diagnostic tests. Examining the proof of Theorem 2 reveals that as *p*_*H*_ increases and *p*_*L*_ decreases, the physicians are more likely to condition their decision on the signal. that is, physicians will only treat high-signal patients with *B* while treating low-signal patients with *N*. Thus, improving the predictability of clinical symptoms or any other available data on the current patient would result in a reduction in the administration of broad-spectrum antibiotics. This conclusion supports Parra-Rodriguez et al. [14] who emphasized the importance of rapid diagnostic tests (RDTs) in the ICU. We showed that these RDTs may help reduce the use of broad-spectrum antibiotics even if they cannot provide certain identification of the pathogen, but merely improve the probabilistic information available for the physician. Hopefully, the increasing development of machine learning tools for clinical decision support systems of antibiotic therapy [37, 38] will facilitate such information flow to physicians.

The gap between the players’ strategic rational behaviour and the socially optimal policy is expected to be particularly pronounced when a large number of patients are to be treated by many independent physicians, each of them making separate and independent clinical decisions. This problem is inherent to the decentralization of decision making, which is a common practice in medicine. A key challenge under these conditions is striking a balance between centralized decision making and the physician’s autonomy. One possible direction is to employ methods of collaborative management and consensus-based regulation of antibiotic prescribing[10]. Another recommended direction is the uptake and integration of decision support systems[14], which combine the patient’s personal data with environment and population-level and data, and thus improve antibiotic choices and help implementing socially optimal considerations.

Nevertheless, before implementing any measures intended to decrease antibiotic misuse, the management or regulator who chooses and designs these measures must take into consideration that physicians are rational players. As such, they are likely to behave in accordance with their utilities, given the new “rules of the game”. It is difficult to impose the socially optimal policy. Instead, the structure of the new game should be set in such a way that the desired behaviour will also be individually rational.

## Data Availability

All data produced in the present work are contained in the manuscript.

## Acknowledgements

The authors would like to thank Professor Dov Samet and Professor Moshe Leshno for their contributions to the long process leading to the final work presented here.

## Funding

This study was supported by the Israel Science Foundation (ISF 1286/21).

## Notes

### Competing Interest Statement

The authors have declared no competing interest.

