## Supplemental Material for "The straight and narrow: a game theory model of broad- and narrow-spectrum empiric antibiotic therapy"

Formulations and proofs of all the theorems, Lemmas and claims.

## SM.1

**Lemma 1.** $d_i^{t,k} = \tilde{BN}, \forall i \in \{1, ..., n\}$ *is a symmetric stage-equilibrium if*

$$\begin{aligned}
&\left(e_B^{t,k} - e_N^{t,k}\right) p_L \\
&\leq \sum_{m=0}^{n-1} \binom{n-1}{m} q_H^m (1-q_H)^{n-1-m} \Big[ v_i^{t+1,k+m} \left(s^{t+1}, \left(d_i^{t,k+m}, d_i^{t,k+m}\right)\right) \\
&- v_i^{t+1,k+m+1} \left(s^{t+1}, \left(d_i^{t,k+m+1}, d_i^{t,k+m+1}\right)\right) \Big] \\
&\leq \left(e_B^{t,k} - e_N^{t,k}\right) p_H
\end{aligned}$$

*Proof.* By the "One-Stage Deviation Principle"[39], it is sufficient to verify that player $i$ cannot gain by deviating from $s$ in a single E-state.

**(i)** In E-state $t, k$ player $i$ has no incentive to deviate to $d_i^{t,k} = \tilde{N}$ if

$$v_i^{t,k}\left(s^{t+1}, \left(\tilde{BN}, \tilde{BN}\right)\right) - v_i^{t,k}\left(s^{t+1}, \left(\tilde{N}, \tilde{BN}\right)\right) \geq 0 \tag{A.1}$$

$$\begin{aligned}
&v_i^{t,k}\left(s^{t+1}, \left(\tilde{BN}, \tilde{BN}\right)\right) - v_i^{t,k}\left(s^{t+1}, \left(\tilde{N}, \tilde{BN}\right)\right) \\
&= \Bigg( \left[ e_B^{t,k} q_H p_H + e_N^{t,k} (1-q_H) p_L \right] \\
&+ \sum_{m=0}^{n} \binom{n}{m} q_H^m (1-q_H)^{n-m} v_i^{t+1,k+m} \left(s^{t+1}, \left(d_i^{t,k+m}, d_i^{t,k+m}\right)\right) \Bigg) \\
&- \Bigg( e_N^{t,k} \left[q_H p_H + (1-q_H) p_L\right] \\
&+ \sum_{m=0}^{n-1} \binom{n-1}{m} q_H^m (1-q_H)^{n-1-m} v_i^{t+1,k+m} \left(s^{t+1}, \left(d_i^{t,k+m}, d_i^{t,k+m}\right)\right) \Bigg) \\
&= \left(e_B^{t,k} - e_N^{t,k}\right) q_H p_H \\
&+ \sum_{m=0}^{n} q_H^m (1-q_H)^{n-1-m} \left[ \binom{n}{m} (1-q_H) - \binom{n-1}{m} \right] v_i^{t+1,k+m} \left(s^{t+1}, \left(d_i^{t,k+m}, d_i^{t,k+m}\right)\right) \\
&= \left(e_B^{t,k} - e_N^{t,k}\right) q_H p_H \\
&+ \sum_{m=0}^{n} q_H \cdot q_H^{m-1} (1-q_H)^{n-1-m} \left[ \left( \binom{n}{m} - \binom{n-1}{m} \right) - \binom{n}{m} q_H \right] \\
&\cdot v_i^{t+1,k+m} \left(s^{t+1}, \left(d_i^{t,k+m}, d_i^{t,k+m}\right)\right)
\end{aligned}$$

Since $\binom{n}{m} = \binom{n-1}{m} + \binom{n-1}{m-1}$ we get

$$
\begin{aligned}
&\left(e_B^{t,k}-e_N^{t,k}\right)q_H p_H\\
&-q_H\sum_{m=0}^{n} q_H^{m-1}\left(1-q_H\right)^{n-1-m}\left[\binom{n}{m}q_H-\binom{n-1}{m-1}\right]v_i^{t+1,k+m}\left(s^{t+1},\left(d_i^{t,k+m},d_i^{t,k+m}\right)\right)\\
&=\left(e_B^{t,k}-e_N^{t,k}\right)q_H p_H\\
&-q_H\sum_{m=0}^{n}\left[\binom{n}{m}q_H^{m}\left(1-q_H\right)^{n-1-m}-\binom{n-1}{m-1}q_H^{m-1}\left(1-q_H\right)^{n-1-m}\right]\\
&\cdot v_i^{t+1,k+m}\left(s^{t+1},\left(d_i^{t,k+m},d_i^{t,k+m}\right)\right)\\
&=\left(e_B^{t,k}-e_N^{t,k}\right)q_H p_H\\
&-q_H\sum_{m=0}^{n}\left[\left(\binom{n-1}{m}+\binom{n-1}{m-1}\right)q_H^{m}\left(1-q_H\right)^{n-1-m}-\binom{n-1}{m-1}q_H^{m-1}\left(1-q_H\right)^{n-1-m}\right]\\
&\cdot v_i^{t+1,k+m}\left(s^{t+1},\left(d_i^{t,k+m},d_i^{t,k+m}\right)\right)\\
&=\left(e_B^{t,k}-e_N^{t,k}\right)q_H p_H\\
&-q_H\sum_{m=0}^{n}\left[\binom{n-1}{m}q_H^{m}\left(1-q_H\right)^{n-1-m}-\binom{n-1}{m-1}q_H^{m-1}\left(1-q_H\right)^{n-1-m}\left(-q_H+1\right)\right]\\
&\cdot v_i^{t+1,k+m}\left(s^{t+1},\left(d_i^{t,k+m},d_i^{t,k+m}\right)\right)\\
&=\left(e_B^{t,k}-e_N^{t,k}\right)q_H p_H\\
&-q_H\sum_{m=0}^{n}\left[\binom{n-1}{m}q_H^{m}\left(1-q_H\right)^{n-1-m}-\binom{n-1}{m-1}q_H^{m-1}\left(1-q_H\right)^{n-m}\right]\\
&\cdot v_i^{t+1,k+m}\left(s^{t+1},\left(d_i^{t,k+m},d_i^{t,k+m}\right)\right)\\
&=\left(e_B^{t,k}-e_N^{t,k}\right)q_H p_H\\
&-q_H\sum_{m=0}^{n-1}\binom{n-1}{m}q_H^{m}\left(1-q_H\right)^{n-1-m}\Big[v_i^{t+1,k+m}\left(s^{t+1},\left(d_i^{t,k+m},d_i^{t,k+m}\right)\right)\\
&-v_i^{t+1,k+m+1}\left(s^{t+1},\left(d_i^{t,k+m+1},d_i^{t,k+m+1}\right)\right)\Big]\\
&\geq 0
\end{aligned}
\tag{A.2}
$$

which yields

$$
\begin{aligned}
&\left(e_B^{t,k}-e_N^{t,k}\right)p_H\\
&\geq\sum_{m=0}^{n-1}\binom{n-1}{m}q_H^{m}\left(1-q_H\right)^{n-1-m}\Big[v_i^{t+1,k+m}\left(s^{t+1},\left(d_i^{t,k+m},d_i^{t,k+m}\right)\right)\\
&-v_i^{t+1,k+m+1}\left(s^{t+1},\left(d_i^{t,k+m+1},d_i^{t,k+m+1}\right)\right)\Big]
\end{aligned}
\tag{A.3}
$$

**(ii)** In E-state $t, k$, Player $i$ has no incentive to deviate to $d_i^{t,k} = \tilde{B}$ if

$$v_i^{t,k}\left(s^{t+1}, \left(\tilde{BN}, \tilde{BN}\right)\right) - v_i^{t,k}\left(s^{t+1}, \left(\tilde{B}, \tilde{BN}\right)\right) \geq 0 \tag{A.4}$$

$$\begin{aligned}
&v_i^{t,k}\left(s^{t+1}, \left(\tilde{BN}, \tilde{BN}\right)\right) - v_i^{t,k}\left(s^{t+1}, \left(\tilde{B}, \tilde{BN}\right)\right) \\
&= \Bigg( \left[e_B^{t,k} q_H p_H + e_N^{t,k}(1-q_H)\, p_L\right] \\
&+ \sum_{m=0}^{n} \binom{n}{m} q_H^m (1-q_H)^{n-m} v_i^{t+1,k+m}\left(s^{t+1}, \left(d_i^{t,k+m}, d_i^{t,k+m}\right)\right) \Bigg) \\
&- \Bigg( e_B^{t,k}\left[q_H p_H + (1-q_H)\, p_L\right] \\
&+ \sum_{m=0}^{n-1} \binom{n-1}{m} q_H^m (1-q_H)^{n-1-m} v_i^{t+1,k+m+1}\left(s^{t+1}, \left(d_i^{t,k+m+1}, d_i^{t,k+m+1}\right)\right) \Bigg) \\
&= \left(e_N^{t,k} - e_B^{t,k}\right)(1-q_H)\, p_L \\
&+ (1-q_H) \sum_{m=0}^{n} q_H^{m-1} (1-q_H)^{n-m-1} \left[\binom{n}{m} q_H - \binom{n-1}{m-1}\right] v_i^{t+1,k+m}\left(s^{t+1}, \left(d_i^{t,k+m}, d_i^{t,k+m}\right)\right) \\
&= \left(e_N^{t,k} - e_B^{t,k}\right)(1-q_H)\, p_L \\
&+ (1-q_H) \sum_{m=0}^{n} \left[\binom{n}{m} q_H^m (1-q_H)^{n-1-m} - \binom{n-1}{m-1} q_H^{m-1} (1-q_H)^{n-1-m}\right] \\
&\cdot v_i^{t+1,k+m}\left(s^{t+1}, \left(d_i^{t,k+m}, d_i^{t,k+m}\right)\right) \\
&\geq 0
\end{aligned} \tag{A.5}$$

which yields

$$\begin{aligned}
&\left(e_B^{t,k} - e_N^{t,k}\right) p_L \\
&\leq \sum_{m=0}^{n-1} \binom{n-1}{m} q_H^m (1-q_H)^{n-1-m} \Big[ v_i^{t+1,k+m}\left(s^{t+1}, \left(d_i^{t,k+m}, d_i^{t,k+m}\right)\right) \\
&- v_i^{t+1,k+m+1}\left(s^{t+1}, \left(d_i^{t,k+m+1}, d_i^{t,k+m+1}\right)\right) \Big]
\end{aligned} \tag{A.6}$$

□

## SM.2

**Lemma 2.** *$d_i^{t,k} = \tilde{BN}, \forall i \in \{1,...,n\}$ is a symmetric stage-optimum if the following two conditions hold:*

$$\begin{aligned}
&\left(e_B^{t,k} - e_N^{t,k}\right)(1-q_H)\,p_L \\
&\leq \sum_{m=0}^{n} \binom{n}{m} q_H^m (1-q_H)^{n-m} \left[ v_i^{t+1,k+m}\left(s^{t+1}, \left(d_i^{t,k+m}, d_i^{t,k+m}\right)\right) \right. \\
&\left. - v_i^{t+1,k+n}\left(s^{t+1}, \left(d_i^{t,k+n}, d_i^{t,k+n}\right)\right) \right]
\end{aligned}$$

*and*

$$\begin{aligned}
&\left(e_B^{t,k} - e_N^{t,k}\right) q_H p_H \\
&\geq \sum_{m=0}^{n} \binom{n}{m} q_H^m (1-q_H)^{n-m} \left[ v_i^{t+1,k}\left(s^{t+1}, \left(d_i^{t,k}, d_i^{t,k}\right)\right) \right. \\
&\left. - v_i^{t+1,k+m}\left(s^{t+1}, \left(d_i^{t,k+m}, d_i^{t,k+m}\right)\right) \right]
\end{aligned}$$

*Proof.* **(i)** In E-state $t, k$ the social planner has no incentive to deviate to $d^{t,k} = \tilde{N}$ if

$$v^{t,k}\left(s^{t+1}, \tilde{BN}\right) - v^{t,k}\left(s^{t+1}, \tilde{N}\right) \geq 0 \tag{A.7}$$

By (13) this condition is equivalent to

$$v_i^{t,k}\left(s^{t+1}, \left(\tilde{BN}, \tilde{BN}\right)\right) - v_i^{t,k}\left(s^{t+1}, \left(\tilde{N}, \tilde{N}\right)\right) \geq 0 \tag{A.8}$$

$$\begin{aligned}
&v_i^{t,k}\left(s^{t+1}, \left(\tilde{BN}, \tilde{BN}\right)\right) - v_i^{t,k}\left(s^{t+1}, \left(\tilde{N}, \tilde{N}\right)\right) \\
&\quad = \Bigg( \left[ e_B^{t,k} q_H p_H + e_N^{t,k} (1-q_H)\, p_L \right] \\
&\quad + \sum_{m=0}^{n} \binom{n}{m} q_H^m (1-q_H)^{n-m} v_i^{t+1,k+m}\left(s^{t+1}, \left(d_i^{t,k+m}, d_i^{t,k+m}\right)\right) \Bigg) \\
&\quad - \Bigg( e_N^{t,k} \left[ q_H p_H + (1-q_H)\, p_L \right] \\
&\quad + v_i^{t+1,k}\left(s^{t+1}, \left(d_i^{t,k}, d_i^{t,k}\right)\right) \Bigg) \\
&\quad = \left(e_B^{t,k} - e_N^{t,k}\right) q_H p_H \\
&\quad + \sum_{m=0}^{n} \binom{n}{m} q_H^m (1-q_H)^{n-m} v_i^{t+1,k+m}\left(s^{t+1}, \left(d_i^{t,k+m}, d_i^{t,k+m}\right)\right) \\
&\quad - v_i^{t+1,k}\left(s^{t+1}, \left(d_i^{t,k}, d_i^{t,k}\right)\right) \geq 0
\end{aligned}$$

which implies

$$\begin{aligned}&\left(e_B^{t,k}-e_N^{t,k}\right)q_Hp_H\\&\geq\sum_{m=0}^{n}\binom{n}{m}q_H^m\left(1-q_H\right)^{n-m}\left[v_i^{t+1,k}\left(s^{t+1},\left(d_i^{t,k},d_i^{t,k}\right)\right)\right.\\&\left.-v_i^{t+1,k+m}\left(s^{t+1},\left(d_i^{t,k+m},d_i^{t,k+m}\right)\right)\right]\end{aligned}\tag{A.9}$$

**(ii)** In E-state $t, k$ the social planner has no incentive to deviate to $d^{t,k} = \tilde{B}$ if

$$v^{t,k}\left(s^{t+1},\tilde{BN}\right)-v^{t,k}\left(s^{t+1},\tilde{B}\right)\geq 0\tag{A.10}$$

By (13) this condition is equivalent to

$$v_i^{t,k}\left(s^{t+1},\left(\tilde{BN},\tilde{BN}\right)\right)-v_i^{t,k}\left(s^{t+1},\left(\tilde{B},\tilde{B}\right)\right)\geq 0\tag{A.11}$$

$$\begin{aligned}&v_i^{t,k}\left(s^{t+1},\left(\tilde{BN},\tilde{BN}\right)\right)-v_i^{t,k}\left(s^{t+1},\left(\tilde{B},\tilde{B}\right)\right)\\&=\left(\left[e_B^{t,k}q_Hp_H+e_N^{t,k}\left(1-q_H\right)p_L\right]\right.\\&\left.+\sum_{m=0}^{n}\binom{n}{m}q_H^m\left(1-q_H\right)^{n-m}v_i^{t+1,k+m}\left(s^{t+1},\left(d_i^{t,k+m},d_i^{t,k+m}\right)\right)\right)\\&-\left(e_B^{t,k}\left[q_Hp_H+\left(1-q_H\right)p_L\right]\right.\\&\left.+v_i^{t+1,k+n}\left(s^{t+1},\left(d_i^{t,k+n},d_i^{t,k+n}\right)\right)\right)\\&=\left(e_N^{t,k}-e_B^{t,k}\right)\left(1-q_H\right)p_L\\&+\sum_{m=0}^{n}\binom{n}{m}q_H^m\left(1-q_H\right)^{n-m}\left[v_i^{t+1,k+m}\left(s^{t+1},\left(d_i^{t,k+m},d_i^{t,k+m}\right)\right)\right.\\&\left.-v_i^{t+1,k+n}\left(s^{t+1},\left(d_i^{t,k+n},d_i^{t,k+n}\right)\right)\right]\geq 0\end{aligned}\tag{A.12}$$

which implies

$$\begin{aligned}&\left(e_B^{t,k}-e_N^{t,k}\right)\left(1-q_H\right)p_L\\&\leq\sum_{m=0}^{n}\binom{n}{m}q_H^m\left(1-q_H\right)^{n-m}\left[v_i^{t+1,k+m}\left(s^{t+1},\left(d_i^{t,k+m},d_i^{t,k+m}\right)\right)\right.\\&\left.-v_i^{t+1,k+n}\left(s^{t+1},\left(d_i^{t,k+n},d_i^{t,k+n}\right)\right)\right]\end{aligned}\tag{A.13}$$

□

## SM.3

**Theorem 1.** *If condition 3 holds then if the decision rule $\tilde{BN}$ at any specific game-tree node (E-state) is part of an MPE, then it is socially optimal at this E-state as well; however, if is socially optimal at any E-state, individual player may have an incentive to deviate towards an excessive use of $B$.*

*Proof.* By lemma 1, in E-state $t, k$, when $d_i^{t,k} = \tilde{BN}, \forall i \in \{1, ..., n\}$, player $i$ has no incentive to deviate to $d_i^{t,k} = \tilde{B}$ if

$$\begin{aligned}&\left(e_B^{t,k} - e_N^{t,k}\right) p_L \\ &\leq \sum_{m=0}^{n-1} \binom{n-1}{m} q_H^m \left(1 - q_H\right)^{n-1-m} \left[v_i^{t+1,k+m} \left(s^{t+1}, \left(d_i^{t,k+m}, d_i^{t,k+m}\right)\right)\right. \\ &\left. - v_i^{t+1,k+m+1} \left(s^{t+1}, \left(d_i^{t,k+m+1}, d_i^{t,k+m+1}\right)\right)\right]\end{aligned} \tag{A.14}$$

And by lemma 2, in E-state $t, k$, when $d_i^{t,k} = \tilde{BN}, \forall i \in \{1, ..., n\}$, the social planner has no incentive to deviate to $d^{t,k} = \tilde{B}, \forall i \in \{1, ..., n\}$ if

$$\begin{aligned}&\left(e_B^{t,k} - e_N^{t,k}\right) \left(1 - q_H\right) p_L \\ &\leq \sum_{m=0}^{n} \binom{n}{m} q_H^m \left(1 - q_H\right)^{n-m} \left[v_i^{t+1,k+m} \left(s^{t+1}, \left(d_i^{t,k+m}, d_i^{t,k+m}\right)\right)\right. \\ &\left. - v_i^{t+1,k+n} \left(s^{t+1}, \left(d_i^{t,k+n}, d_i^{t,k+n}\right)\right)\right]\end{aligned} \tag{A.15}$$

$$\begin{aligned}&\sum_{m=0}^{n} \binom{n}{m} q_H^m \left(1 - q_H\right)^{n-m} \left[v_i^{t+1,k+m} \left(s^{t+1}, \left(d_i^{t,k+m}, d_i^{t,k+m}\right)\right)\right. \\ &\left. - v_i^{t+1,k+n} \left(s^{t+1}, \left(d_i^{t,k+n}, d_i^{t,k+n}\right)\right)\right] \\ &= \sum_{m=0}^{n-1} \binom{n}{m} q_H^m \left(1 - q_H\right)^{n-m} \left[v_i^{t+1,k+m} \left(s^{t+1}, \left(d_i^{t,k+m}, d_i^{t,k+m}\right)\right)\right. \\ &\left. - v_i^{t+1,k+n} \left(s^{t+1}, \left(d_i^{t,k+n}, d_i^{t,k+n}\right)\right)\right] \\ &= \left(1 - q_H\right) \sum_{m=0}^{n-1} \binom{n}{m} q_H^m \left(1 - q_H\right)^{n-m-1} \left[v_i^{t+1,k+m} \left(s^{t+1}, \left(d_i^{t,k+m}, d_i^{t,k+m}\right)\right)\right. \\ &\left. - v_i^{t+1,k+n} \left(s^{t+1}, \left(d_i^{t,k+n}, d_i^{t,k+n}\right)\right)\right]\end{aligned}$$

And therefore (A.15) can be presented as

$$\begin{aligned}&\left(e_B^{t,k} - e_N^{t,k}\right) p_L \\ &\leq \sum_{m=0}^{n-1} \binom{n}{m} q_H^m \left(1 - q_H\right)^{n-m-1} \left[v_i^{t+1,k+m} \left(s^{t+1}, \left(d_i^{t,k+m}, d_i^{t,k+m}\right)\right)\right. \\ &\left. - v_i^{t+1,k+n} \left(s^{t+1}, \left(d_i^{t,k+n}, d_i^{t,k+n}\right)\right)\right]\end{aligned} \tag{A.16}$$

Comparing (A.16) and (A.14)

$$\begin{aligned}
&\sum_{m=0}^{n-1}\binom{n}{m}q_H^m\left(1-q_H\right)^{n-1-m}\bigg[v_i^{t+1,k+m}\left(s^{t+1},\left(d_i^{t,k+m},d_i^{t,k+m}\right)\right)\\
&-v_i^{t+1,k+n}\left(s^{t+1},\left(d_i^{t,k+n},d_i^{t,k+n}\right)\right)\bigg]\\
&-\sum_{m=0}^{n-1}\binom{n-1}{m}q_H^m\left(1-q_H\right)^{n-1-m}\bigg[v_i^{t+1,k+m}\left(s^{t+1},\left(d_i^{t,k+m},d_i^{t,k+m}\right)\right)\\
&-v_i^{t+1,k+m+1}\left(s^{t+1},\left(d_i^{t,k+m+1},d_i^{t,k+m+1}\right)\right)\bigg]\\
&=\sum_{m=0}^{n-1}\binom{n-1}{m-1}q_H^m\left(1-q_H\right)^{n-1-m}\bigg[v_i^{t+1,k+m}\left(s^{t+1},\left(d_i^{t,k+m},d_i^{t,k+m}\right)\right)\\
&-v_i^{t+1,k+m+1}\left(s^{t+1},\left(d_i^{t,k+m+1},d_i^{t,k+m+1}\right)\right)\bigg]\\
&+\sum_{m=0}^{n-1}\binom{n}{m}q_H^m\left(1-q_H\right)^{n-1-m}\bigg[v_i^{t+1,k+m+1}\left(s^{t+1},\left(d_i^{t,k+m+1},d_i^{t,k+m+1}\right)\right)\\
&-v_i^{t+1,k+n}\left(s^{t+1},\left(d_i^{t,k+n},d_i^{t,k+n}\right)\right)\bigg]
\end{aligned}\tag{A.17}$$

If condition 3 holds then both addends are positive. Therefore, (A.14) implies (A.16), but it is possible that (A.16) would hold and (A.14) would not. Meaning, if $d_i^{t,k}=\tilde{BN},\forall i\in\{1,...,n\}$ is a stage-equilibrium then it is necessarily a stage-social-optimum as well; however, it is possible that $d_i^{t,k}=\tilde{BN},\forall i\in\{1,...,n\}$ is a stage-social-optimum, but (each) player $i$ has an incentive to unilaterally deviate to $d_i^{t,k}=\tilde{B}$.

Similarly, it can be shown that if $d_i^{t,k}=\tilde{N},\forall i\in\{1,...,n\}$ is a stage-equilibrium then it is necessarily a stage-social-optimum as well; however, it is possible that $d_i^{t,k}=\tilde{N},\forall i\in\{1,...,n\}$ is a stage-social-optimum, but (each) player $i$ has an incentive to unilaterally deviate to $d_i^{t,k}=\tilde{B}$.

□

## SM.4

**Claim 1.** *In MPE and in the socially optimal policy,*

$$d_i^{T,k}=\tilde{B},\forall k\in\{0,1,...,nT\},\forall i\in\{1,...,n\}$$

*Proof.* In period $T$, the expected payoff of player $i$ in E-state $T,k$ is:

$$v_i^{T,k}\left(d_i^{T,k},d_{-i}^{T,k}\right)=\sum_{j\in\{L,H\}}\left[P\left(B\middle|j,d_i^{T,k}\right)q_jp_je_B^{T,k}+P\left(N\middle|j,d_i^{T,k}\right)q_jp_je_N^{T,k}\right]\tag{A.18}$$

i.e.

$$\begin{aligned}
v_i^{T,k}\left(\tilde{B},d_{-i}^{T,k}\right)&=e_B^{T,k}\left[q_Hp_H+\left(1-q_H\right)p_L\right]\\
v_i^{T,k}\left(\tilde{BN},d_{-i}^{T,k}\right)&=e_B^{T,k}q_Hp_H+e_N^{T,k}\left(1-q_H\right)p_L\\
v_i^{T,k}\left(\tilde{N},d_{-i}^{T,k}\right)&=e_N^{T,k}\left[q_Hp_H+\left(1-q_H\right)p_L\right]
\end{aligned}\tag{A.19}$$

And because (4)

$$e_B^{T,k}>e_N^{T,k}\forall k\in\{0,1,...,nT\}\tag{A.20}$$

we get that

$$v_i^{T,k}\left(\tilde{B}, d_{-i}^{T,k}\right) > v_i^{T,k}\left(\tilde{BN}, d_{-i}^{T,k}\right) > v_i^{T,k}\left(\tilde{N}, d_{-i}^{T,k}\right) \tag{A.21}$$

□

## SM.5

**Claim 2.** *In a fixed symmetric strategy profile where* $d_i^{t,k} = \tilde{BN}, \forall i \in \{1,...,n\}, \forall k \in \{0,1,...,n(T-1)\}, t = 0,...,T-1$ *and* $d_i^{T,k} = \tilde{B}, \forall i \in \{1,...,n\}, \forall k \in \{0,1,...,nT\}$

$$\begin{aligned} & v_i^{t,k}\left(s^{t+1}, \left(d_i^{t,k}, d_i^{t,k}\right)\right) - v_i^{t,k+1}\left(s^{t+1}, \left(d_i^{t,k+1}, d_i^{t,k+1}\right)\right) \\ & \quad = (T-t)\left[\alpha_B q_H p_H - I\alpha_N \left(1-q_H\right) p_L\right] + \alpha_B \left[q_H p_H + \left(1-q_H\right) p_L\right] \end{aligned} \tag{A.22}$$

*Proof.* By induction.

(A.22) holds for $t = T$:
By claim 1, in period $T$, the expected payoff of player $i$ in E-state $T,k$ is:

$$v_i^{T,k}\left(\tilde{B}, \tilde{B}\right) = e_B^{T,k}\left[q_H p_H + \left(1-q_H\right) p_L\right] \tag{A.23}$$

Therefore for $t = T$,

$$\begin{aligned} & v_i^{t,k}\left(s^{t+1}, \left(d_i^{t,k}, d_i^{t,k}\right)\right) - v_i^{t,k+1}\left(s^{t+1}, \left(d_i^{t,k+1}, d_i^{t,k+1}\right)\right) \\ & \quad = v_i^{T,k}\left(\tilde{B}, \tilde{B}\right) - v_i^{T,k+1}\left(\tilde{B}, \tilde{B}\right) \\ & \quad = e_B^{T,k}\left[q_H p_H + \left(1-q_H\right) p_L\right] - e_B^{T,k+1}\left[q_H p_H + \left(1-q_H\right) p_L\right] \\ & \quad = \alpha_B \left[q_H p_H + \left(1-q_H\right) p_L\right] \\ & \quad = (T-T)\left[\alpha_B q_H p_H - I\alpha_N \left(1-q_H\right) p_L\right] + \alpha_B \left[q_H p_H + \left(1-q_H\right) p_L\right] \end{aligned} \tag{A.24}$$

Assume that (A.22) holds for some $t+1$; then for a fixed symmetric strategy profile where $d_i^{t,k} = \tilde{BN}, \forall k \in \{0,1,...,nt\}, \forall i \in \{1,...,n\}$ it holds for $t$ as well:

$$
\begin{aligned}
& v_i^{t,k}\left(s^{t+1},\left(\tilde{BN},\tilde{BN}\right)\right)-v_i^{t,k+1}\left(s^{t+1},\left(\tilde{BN},\tilde{BN}\right)\right) \\
&= \Bigg(\left[e_B^{t,k}q_Hp_H+e_N^{t,k}\left(1-q_H\right)p_L\right] \\
&+\sum_{m=0}^{n}\binom{n}{m}q_H^m\left(1-q_H\right)^{n-m}v_i^{t+1,k+m}\left(s^{t+1},\left(d_i^{t,k+m},d_i^{t,k+m}\right)\right)\Bigg) \\
&-\Bigg(\left[e_B^{t,k+1}q_Hp_H+e_N^{t,k+1}\left(1-q_H\right)p_L\right] \\
&+\sum_{m=0}^{n}\binom{n}{m}q_H^m\left(1-q_H\right)^{n-m}v_i^{t+1,k+m+1}\left(s^{t+1},\left(d_i^{t,k+m+1},d_i^{t,k+m+1}\right)\right)\Bigg) \\
&=\alpha_Bq_Hp_H-I\alpha_N\left(1-q_H\right)p_L \\
&+\sum_{m=0}^{n}\binom{n}{m}q_H^m\left(1-q_H\right)^{n-m}\Bigg[v_i^{t+1,k+m}\left(s^{t+2},\left(d_i^{t+1,k+m},d_i^{t+1,k+m}\right)\right) \\
&-v_i^{t+1,k+m+1}\left(s^{t+2},\left(d_i^{t+1,k+m+1},d_i^{t+1,k+m+1}\right)\right)\Bigg] \\
&=\alpha_Bq_Hp_H-I\alpha_N\left(1-q_H\right)p_L \\
&+\sum_{m=0}^{n}\binom{n}{m}q_H^m\left(1-q_H\right)^{n-m}\Bigg[\left(T-(t+1)\right)\left[\alpha_Bq_Hp_H-\alpha_N\left(1-q_H\right)p_L\right] \\
&+\alpha_B\left[q_Hp_H+\left(1-q_H\right)p_L\right]\Bigg] \\
&=\alpha_Bq_Hp_H-I\alpha_N\left(1-q_H\right)p_L \\
&+\big[\left(T-t-1\right)\left[\alpha_Bq_Hp_H-I\alpha_N\left(1-q_H\right)p_L\right]+\alpha_B\left[q_Hp_H+\left(1-q_H\right)p_L\right]\big] \\
&=\left(T-t\right)\left[\alpha_Bq_Hp_H-I\alpha_N\left(1-q_H\right)p_L\right]+\alpha_B\left[q_Hp_H+\left(1-q_H\right)p_L\right]
\end{aligned} \tag{A.25}
$$

□

## SM.6

**Lemma 3.** *The fixed symmetric strategy profile where* $d_i^{t,k}=\tilde{BN},\forall k\in\{0,1,...,nt\},\forall i\in\{1,...,n\},t=0,...,T-1$ *and* $d_i^{T,k}=\tilde{B},\forall k\in\{0,1,...,nT\},\forall i\in\{1,...,n\}$ *is a MPE if*

$$
\begin{aligned}
&\left(e_B^{t,k}-e_N^{t,k}\right)p_L \\
&\leq\left[T-(t+1)\right]\left[\alpha_Bq_Hp_H-I\alpha_N\left(1-q_H\right)p_L\right]+\alpha_B\left[q_Hp_H+\left(1-q_H\right)p_L\right] \\
&\leq\left(e_B^{t,k}-e_N^{t,k}\right)p_H
\end{aligned}
$$

*Proof.* By the "One-Stage Deviation Principle"[], it is sufficient to verify that player $i$ cannot gain by deviating from $s$ in a single E-state.

**(i)** By lemma 1, in E-state $t,k$ player $i$ has no incentive to deviate to $d_i^{t,k}=\tilde{N}$ if

$$
\begin{aligned}
&\left(e_B^{t,k}-e_N^{t,k}\right)p_H \\
&\geq\sum_{m=0}^{n-1}\binom{n-1}{m}q_H^m\left(1-q_H\right)^{n-1-m}\Bigg[v_i^{t+1,k+m}\left(s^{t+1},\left(d_i^{t,k+m},d_i^{t,k+m}\right)\right) \\
&-v_i^{t+1,k+m+1}\left(s^{t+1},\left(d_i^{t,k+m+1},d_i^{t,k+m+1}\right)\right)\Bigg]
\end{aligned} \tag{A.26}
$$

By claim 2:

$$\begin{aligned}&\sum_{m=0}^{n-1}\binom{n-1}{m}q_H^m\left(1-q_H\right)^{n-1-m}\Big[v_i^{t+1,k+m}\left(s^{t+1},\left(d_i^{t,k+m},d_i^{t,k+m}\right)\right)\\&-v_i^{t+1,k+m+1}\left(s^{t+1},\left(d_i^{t,k+m+1},d_i^{t,k+m+1}\right)\right)\Big]\\&=\left(T-(t+1)\right)\left[\alpha_B q_H p_H-I\alpha_N\left(1-q_H\right)p_L\right]+\alpha_B\left[q_H p_H+\left(1-q_H\right)p_L\right]\end{aligned}\tag{A.27}$$

which yields

$$\begin{aligned}&\left(e_B^{t,k}-e_N^{t,k}\right)p_H\\&\geq\left(T-(t+1)\right)\left[\alpha_B q_H p_H-I\alpha_N\left(1-q_H\right)p_L\right]+\alpha_B\left[q_H p_H+\left(1-q_H\right)p_L\right]\end{aligned}\tag{A.28}$$

**(ii)** By lemma 1, in E-state $t,k$, Player $i$ has no incentive to deviate to $d_i^{t,k}=\tilde{B}$ if

$$\begin{aligned}&\left(e_B^{t,k}-e_N^{t,k}\right)p_L\\&\leq\sum_{m=0}^{n-1}\binom{n-1}{m}q_H^m\left(1-q_H\right)^{n-1-m}\Big[v_i^{t+1,k+m}\left(s^{t+1},\left(d_i^{t,k+m},d_i^{t,k+m}\right)\right)\\&-v_i^{t+1,k+m+1}\left(s^{t+1},\left(d_i^{t,k+m+1},d_i^{t,k+m+1}\right)\right)\Big]\end{aligned}\tag{A.29}$$

And by claim 2:

$$\begin{aligned}&\left(e_B^{t,k}-e_N^{t,k}\right)p_L\\&\leq\left(T-(t+1)\right)\left[\alpha_B q_H p_H-I\alpha_N\left(1-q_H\right)p_L\right]+\alpha_B\left[q_H p_H+\left(1-q_H\right)p_L\right]\end{aligned}\tag{A.30}$$

□

## SM.7

**Lemma 4.** *The fixed symmetric strategy profile where $d^{t,k}=\tilde{BN},\forall k\in\{0,1,...,nt\},t=0,...,T-1$ and $d^{T,k}=\tilde{B},\forall k\in\{0,1,...,nT\}$ is an optimal policy if*

$$\begin{aligned}&\left(e_B^{t,k}-e_N^{t,k}\right)p_L\\&\leq n\Big(\left[T-(t+1)\right]\left[\alpha_B q_H p_H-I\alpha_N\left(1-q_H\right)p_L\right]+\alpha_B\left[q_H p_H+\left(1-q_H\right)p_L\right]\Big)\\&\leq\left(e_B^{t,k}-e_N^{t,k}\right)p_H\end{aligned}$$

*Proof.* **(i)** By lemma 2, in E-state $t,k$ the social planner has no incentive to deviate to $d^{t,k}=\tilde{N}$ if

$$\begin{aligned}&\left(e_B^{t,k}-e_N^{t,k}\right)q_H p_H\\&\geq\sum_{m=0}^{n}\binom{n}{m}q_H^m\left(1-q_H\right)^{n-m}\Big[v_i^{t+1,k}\left(s^{t+1},\left(d_i^{t,k},d_i^{t,k}\right)\right)\\&-v_i^{t+1,k+m}\left(s^{t+1},\left(d_i^{t,k+m},d_i^{t,k+m}\right)\right)\Big]\end{aligned}\tag{A.31}$$

Let $\delta v_i^{t+1,k} = v_i^{t+1,k}\left(s^{t+1},\left(d_i^{t,k},d_i^{t,k}\right)\right) - v_i^{t+1,k+1}\left(s^{t+1},\left(d_i^{t,k+1},d_i^{t,k+1}\right)\right)$.

$$\begin{aligned}
&\sum_{m=0}^{n}\binom{n}{m}q_H^m\left(1-q_H\right)^{n-m}\Big[v_i^{t+1,k}\left(s^{t+1},\left(d_i^{t,k},d_i^{t,k}\right)\right)\\
&-v_i^{t+1,k+m}\left(s^{t+1},\left(d_i^{t,k+m},d_i^{t,k+m}\right)\right)\Big]\\
&=\sum_{m=0}^{n}\binom{n}{m}q_H^m\left(1-q_H\right)^{n-m}m\delta v_i^{t+1,k}\\
&=\delta v_i^{t+1,k}\left(\sum_{m=0}^{n}\binom{n}{m}q_H^m\left(1-q_H\right)^{n-m}m\right)\\
&=\delta v_i^{t+1,k}\cdot nq_H
\end{aligned}$$

Therefore, (A.31) and claim 2 implies

$$\begin{aligned}
\left(e_B^{t,k}-e_N^{t,k}\right)p_H &\geq n\delta v_i^{t+1,k}\\
&=n\Big[v_i^{t+1,k}\left(s^{t+1},\left(d_i^{t,k},d_i^{t,k}\right)\right)\\
&-v_i^{t+1,k+1}\left(s^{t+1},\left(d_i^{t,k+1},d_i^{t,k+1}\right)\right)\Big]\\
&=n\Big[\left(T-(t+1)\right)\left[\alpha_B q_H p_H - I\alpha_N\left(1-q_H\right)p_L\right]+\alpha_B\left[q_H p_H+\left(1-q_H\right)p_L\right]\Big]
\end{aligned}\tag{A.32}$$

**(ii)** By lemma 2, in E-state $t,k$ the social planner has no incentive to deviate to $d^{t,k}=\tilde{B}$ if

$$\begin{aligned}
&\left(e_B^{t,k}-e_N^{t,k}\right)\left(1-q_H\right)p_L\\
&\leq\sum_{m=0}^{n}\binom{n}{m}q_H^m\left(1-q_H\right)^{n-m}\Big[v_i^{t+1,k+m}\left(s^{t+1},\left(d_i^{t,k+m},d_i^{t,k+m}\right)\right)\\
&-v_i^{t+1,k+n}\left(s^{t+1},\left(d_i^{t,k+n},d_i^{t,k+n}\right)\right)\Big]
\end{aligned}\tag{A.33}$$

Using $\delta v_i^{t+1,k} = v_i^{t+1,k}\left(s^{t+1},\left(d_i^{t,k},d_i^{t,k}\right)\right) - v_i^{t+1,k+1}\left(s^{t+1},\left(d_i^{t,k+1},d_i^{t,k+1}\right)\right)$.

$$\begin{aligned}
&\sum_{m=0}^{n}\binom{n}{m}q_H^m\left(1-q_H\right)^{n-m}\Big[v_i^{t+1,k+m}\left(s^{t+1},\left(d_i^{t,k+m},d_i^{t,k+m}\right)\right)\\
&-v_i^{t+1,k+n}\left(s^{t+1},\left(d_i^{t,k+n},d_i^{t,k+n}\right)\right)\Big]\\
&=\sum_{m=0}^{n}\binom{n}{m}q_H^m\left(1-q_H\right)^{n-m}\left(n-m\right)\delta v_i^{t+1,k}\\
&=\delta v_i^{t+1,k}\left(n-\sum_{m=0}^{n}\binom{n}{m}q_H^m\left(1-q_H\right)^{n-m}m\right)\\
&=\delta v_i^{t+1,k}\left(n-nq_H\right)\\
&=n\left(1-q_H\right)\delta v_i^{t+1,k}
\end{aligned}$$

Therefore, (A.33) implies

$$
\begin{aligned}
\left(e_B^{t,k} - e_N^{t,k}\right) p_L &\leq n\delta v_i^{t+1,k} \\
&= n\Big[ v_i^{t+1,k}\left(s^{t+1}, \left(d_i^{t,k}, d_i^{t,k}\right)\right) \\
&- v_i^{t+1,k+1}\left(s^{t+1}, \left(d_i^{t,k+1}, d_i^{t,k+1}\right)\right)\Big] \\
&= n\Big[ (T-(t+1))\left[\alpha_B q_H p_H - I\alpha_N (1-q_H) p_L\right] + \alpha_B \left[q_H p_H + (1-q_H) p_L\right]\Big]
\end{aligned}
\tag{A.34}
$$

□

## SM.8

**Theorem 2.** *If condition 3 holds, then if the fixed symmetric strategy profile where* $d_i^{t,k} = \tilde{BN}, \forall k \in \{0,1,...,nt\}, \forall i \in \{1,...,n\}, t = 0,...,T-1$ *and* $d_i^{T,k} = \tilde{B}, \forall k \in \{0,1,...,nT\}, \forall i \in \{1,...,n\}$ *("the signal-based policy") is socially optimal, individual player may have an incentive to deviate towards an excessive use of* $B$*. Furthermore, if this strategy profile is not optimal for a given combination of medical parameters* $(e_B, e_N, \alpha_B, \alpha_N, q_H, p_H, p_L, I)$ *when* $n = 1$*, it may be optimal as the number of patients grows, but will never be an MPE.*

*Proof.* By lemma 3 the fixed symmetric strategy profile where $d_i^{t,k} = \tilde{BN}, \forall k \in \{0,1,...,nt\}, \forall i \in \{1,...,n\}, t = 0,...,T-1$ and $d_i^{T,k} = \tilde{B}, \forall k \in \{0,1,...,nT\}, \forall i \in \{1,...,n\}$ is a MPE if

$$
\begin{aligned}
&\left(e_B^{t,k} - e_N^{t,k}\right) p_L \\
&\leq [T-(t+1)]\left[\alpha_B q_H p_H - I\alpha_N (1-q_H) p_L\right] + \alpha_B \left[q_H p_H + (1-q_H) p_L\right] \\
&\leq \left(e_B^{t,k} - e_N^{t,k}\right) p_H
\end{aligned}
$$

And by lemma 4 the fixed symmetric strategy profile where $d^{t,k} = \tilde{BN}, \forall k \in \{0,1,...,nt\}, t = 0,...,T-1$ and $d^{T,k} = \tilde{B}, \forall k \in \{0,1,...,nT\}$ is an optimal policy if

$$
\begin{aligned}
&\left(e_B^{t,k} - e_N^{t,k}\right) p_L \\
&\leq n\Big( [T-(t+1)]\left[\alpha_B q_H p_H - I\alpha_N (1-q_H) p_L\right] + \alpha_B \left[q_H p_H + (1-q_H) p_L\right]\Big) \\
&\leq \left(e_B^{t,k} - e_N^{t,k}\right) p_H
\end{aligned}
$$

And if condition 3 holds then

$$
\begin{aligned}
&n\Big( [T-(t+1)]\left[\alpha_B q_H p_H - I\alpha_N (1-q_H) p_L\right] + \alpha_B \left[q_H p_H + (1-q_H) p_L\right]\Big) \\
&\geq [T-(t+1)]\left[\alpha_B q_H p_H - I\alpha_N (1-q_H) p_L\right] + \alpha_B \left[q_H p_H + (1-q_H) p_L\right]
\end{aligned}
$$

□
